# Hypnotherapy for major depressive disorder: a systematic review of randomized clinical trials

**DOI:** 10.1101/2024.07.15.24310435

**Authors:** Filipe Luís Souza, Murilo Schiefler Moura, Gary Elkins, Juliana Vieira Almeida Silva

**Affiliations:** School of Health Sciences, Universidade do Vale do Itajaí, Santa Catarina, Brazil; Department of Psychology and Neuroscience, Baylor University, Texas, USA

## Abstract

**Introduction:** Major Depressive Disorder (MDD) is one of the most debilitating diseases worldwide and has seen a significant increase in diagnoses during the pandemic, demanding more and better therapeutic tools to manage the post-pandemic scenario. **Objective:** The aim of this systematic review is to provide a comprehensive analysis of the last 20 years of clinical research on Hypnotherapy (HT) to determine whether this intervention has evidence to support its recommendation for the treatment of MDD. **Methods:** This review included only randomized clinical trials (RCTs) involving adult populations diagnosed with MDD, regardless of the severity level (mild, moderate, or severe) according to any validated diagnostic criteria, compared to a control group (active treatment or none), with any follow-up duration and free access to the manuscript. The bibliographic survey was planned to be as sensitive as possible, conducted across seven distinct databases: MEDLINE (PubMed), Embase, CENTRAL, PsycINFO, Scopus, ScieELO, and Latin American and Caribbean Health Sciences Literature (LILACS). To identify potentially eligible studies in the grey literature, researchers also searched the U.S. National Library of Medicine (ClinicalTrials.gov). The risk of bias was assessed by two independent investigators using Cochrane’s revised tool (RoB 2), and the final judgment was made by consensus. To better analyze the included studies, the certainty of the evidence was evaluated through the Grading of Recommendations Assessment, Development and Evaluation (GRADE). Due to the lack of comparable studies, it was not possible to perform a meta-analysis; therefore, the studies were graphically displayed in Descriptive Forest Plots. **Results:** There is very low-quality evidence suggesting that hypnosis-based interventions may reduce the severity of depression, which precludes the clinical recommendation of this intervention for patients in the real world, pending the production of better evidence of effectiveness and safety, although no evidence of significant adverse effects was found. **Other:** This review was pre-registered with PROSPERO and can be accessed using the registration number CRD42023409631.

## 1. INTRODUCTION

Characterized as an affective mood disorder, Major Depressive Disorder (MDD) is defined by intense sadness lasting at least two weeks, resulting in functional, molecular, and structural changes in various areas of the brain (Bains & Abdijadid, 2023). Among the consequences and comorbidities of MDD, there is a high risk of progression to death by suicide, with an approximate chance of 15% (Orsolini et al., 2020), and suicide is responsible annually for more deaths than malaria, HIV/AIDS, war, homicide, and breast cancer (WHO, 2021). However, it is estimated that the COVID-19 pandemic triggered around 53.2 million new cases of MDD worldwide, particularly affecting women (Santomauro et al., 2021). Given the increasing epidemiological data on Major Depressive Disorder (MDD), there is a clear need and importance for effective, empirically based, and accessible clinical interventions for the treatment of mood disorders and all sorts of mental disorders. An alternative that is receiving increasing attention is hypnotherapy (HT).

Despite hypnosis being the first form of psychotherapy in the West (DeSouza et al., 2020) and the significant accumulation of evidence regarding functional brain changes associated with hypnosis (Wolf et al., 2022), Souza and Broering (2022) identified that the majority of hypnosis research is conducted from a state hypothesis theoretical line, varying between qualitative and quantitative views, weak state, and that many authors make minor alterations to their definitions of the hypnotic phenomenon, creating a spectrum of definitions without empirical foundation. Although most hypnosis research treats it as an altered state of consciousness, this definition of the phenomenon, in over 100 years of research, has not demonstrated a causal relationship between functional brain changes and the human capacity for response to suggestions (Mazzoni et al., 2013) and the identified changes in neural activity patterns. To date, the alleged changes from a special state of consciousness may simply be linked to standard variations in brain activity (Tuominen et al., 2021); nevertheless, this theoretical perspective continues to underpin research and clinical trials in the field.

The broad variability in clinical application methods and the lack of cohesion in defining hypnosis as a phenomenon have driven the creation of guidelines to evaluate the efficacy of clinical hypnosis applications and to conduct new research. These guidelines were developed by a task force comprising researchers from Hungary, Italy, the United Kingdom, and the United States of America (Kekecs et al., 2022). Subsequently, the task force conducted a survey with healthcare professionals from 31 countries who use hypnosis in their clinical practices, ranking hypnosis applications based on their personal experiences and perceptions of the intervention’s effectiveness. Among the 37 applications listed, depression was considered one of the least effective, despite being commonly promoted as an effective target for hypnosis treatment (Palsson et al., 2023), highlighting another relevant factor for conducting this research. Recognizing the challenges in defining hypnosis-based interventions and managing biases resulting from different applications by physicians adept in the technique, the task force incorporated these limitations into the formulation of guidelines to enhance methodological rigor in hypnosis research (Yapko, 2022).

To determine if clinicians around the world now have a new alternative to aid in the treatment of MDD in a post-pandemic scenario, this review seeks to analyze the available evidence in RCTs on the potential reduction of depressive symptoms in individuals with MDD produced by HT. In this study we aim to address the following specific research objectives: O1) selecting RCTs that examined the effects of HT on MDD; O2) evaluating the methodological quality of the selected studies; O3) extracting relevant data from the selected trials; O4: assessing the clinical efficacy outcomes of HT compared to Cognitive Behavioral Therapy (CBT).

## 2. METHODS

Eligibility criteria were divided into two categories: 1) Inclusion criteria: randomized clinical trials, including cluster-randomized trials, conducted within the past 20 years. The RCT must feature a control group (e.g., gold-standard therapy) or a comparison group that did not receive any specific psychological intervention (e.g., waitlist), and must provide individual data from participants diagnosed with depression, in case the research involved a mixed population (also including healthy individuals); studies must be freely accessible; the population must be over 18 years old; diagnosed with MDD, regardless of severity level (mild, moderate, or severe) according to any validated diagnostic criteria, such as the Diagnostic and Statistical Manual of Mental Disorders (DSM), International Classification of Diseases (ICD), or other validated criteria. 2) Exclusion criteria: studies that conducted trials with bipolar depressive patients or other diagnoses; trials with populations under 18 years of age; observational studies, literature reviews, non-randomized clinical trials; clinical trials without a control group comparison; incomplete texts will not be included; comorbid disorders, such as anxiety disorders, are not an exclusion criterion from the trial population as long as the individual has MDD as the primary diagnosis; studies that included participants with neurodegenerative diseases. Inclusion and exclusion criteria were developed from the Population, Intervention, Comparison, Outcome, Timing, and Study Design (PICOTS) method, presented in Table 1.

**Table 1.**
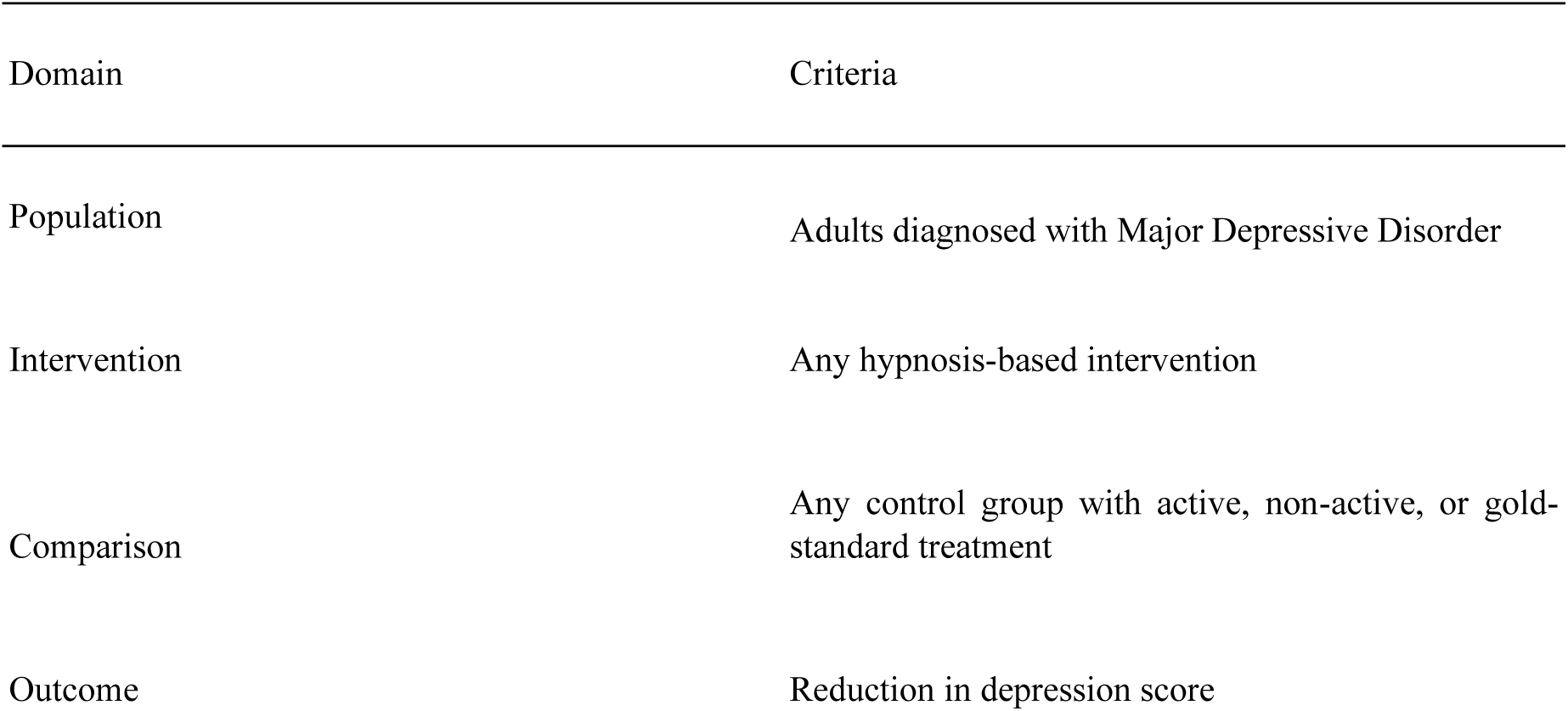
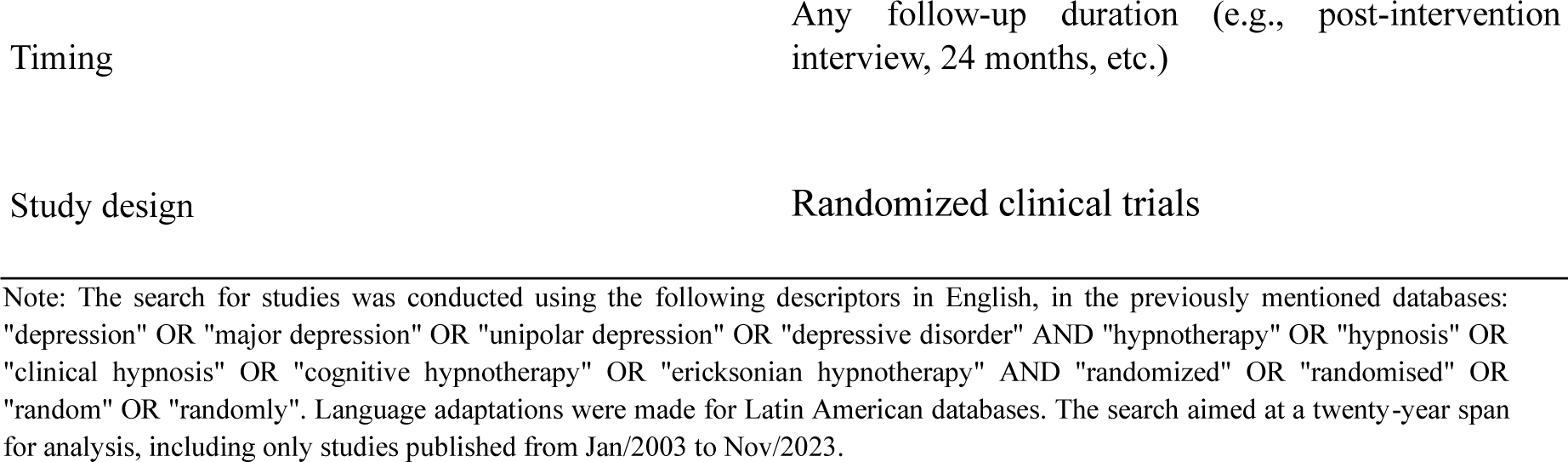
PICOTS Strategy for Comprehensive Inclusion of Studies.

To construct the search strategy and define the databases to be used, two 1-hour meetings were held with a team of librarians from a community university. Searches to identify RCTs were conducted across seven databases: MEDLINE (PubMed), Embase, CENTRAL, PsycINFO, Scopus, ScieELO, and Latin American and Caribbean Health Sciences Literature (LILACS). Records were identified and selected based on their titles and abstracts according to the previously described eligibility criteria by two independent investigators (F.L.S and M.S.M.). A pilot test of the eligibility screening and data extraction was conducted using a single study indicated by another investigator (J.V.A.S.). To identify potentially eligible studies in the grey literature, records were also searched in the U.S. National Library of Medicine (ClinicalTrials.gov). After eliminating studies irrelevant to the research question and duplicates, the full texts of the articles were independently assessed for their eligibility for the review by the investigators. This process was conducted using Covidence software for systematic review reference management. Disagreements between researchers were resolved by consensus.

Data were extracted using the Cochrane Training’s Data Extraction and Assessment Form (<u>Training Cochrane</u>) by two independent investigators (F.L.S and M.S.M.). Data on the primary outcomes assessed (reduction in depression severity) in each included RCT, its direction and timing, were collected. Studies that did not assess the primary outcome in continuous outcomes were not excluded from the review. However, if a study assessed the primary outcome in both dichotomous and continuous outcomes, the data from the continuous outcome were chosen for extraction. This decision was made by consensus among the researchers.

### 2.1 BIAS RISK ASSESSMENT

The risk of bias in the included RCTs was assessed by two independent investigators (F.L.S. and M.S.M) using the Cochrane’s revised tool - RoB 2 (Sterne et al., 2019), and the final judgment was decided by consensus. The investigators receive private training from two researchers experienced in publishing Cochrane systematic reviews. The biases of the selected studies were evaluated based on the following criteria: 1) bias arising from the randomization process; 2) bias due to deviations from intended interventions; 3) bias due to missing outcome data; 4) bias in outcome measurement; 5) bias in the selection of the reported outcome. After the individual analysis by the investigators, the final risk of bias judgment will be: A) low risk of bias; B) some concerns; and C) high risk of bias.

### 2.2 TREATMENT EFFECT MEASURES

#### Dichotomous Outcomes

Dichotomous data were analyzed using Odds Ratio (OR) and 95% Confidence Intervals (CI) for each effect estimate.

#### Continuous Outcomes

Continuous data presented in Descriptive Forest Plots were analyzed using Mean Differences (MD) and confidence intervals calculated at 95%. To discuss limitations of the evidence found, a simulated meta-analysis was conducted using Standardized Mean Differences (SMD) to combine different scales of effect measurement into a single analysis and with a random effects analysis due to the heterogeneity of the studies included for hypothetical analysis (see supplementary materials). These syntheses were generated using Review Manager 5.4.

### 2.3 ASSESSMENT OF EVIDENCE CERTAINTY

To better analyze the evidence presented in the selected studies, this review utilized the GRADE approach to recommend or not recommend HT for the treatment of MDD. For each PICOT generated from the included studies, the quality of evidence was initially considered “high” and subsequently downgraded to “moderate,” “low,” or “very low” based on the following criteria: 1) risk of bias within the study; 2) inconsistency; 3) indirect evidence; 4) imprecision; and 5) risk of publication bias.

## 3. RESULTS

A systematic search was designed to have higher sensitivity in the literature search, increasing the likelihood of finding more eligible studies, as well as the number of studies classified as duplicates (N = 351), removed manually and also by reference manager. Not excluding works based on their language, initially, 4411 studies were found (see Figure 1). More than half of the studies were removed for not being of the appropriate study design (N = 1156) or for studying another population (N = 1213), e.g., cancer, pain, children, etc. Leaving potentially eligible studies (N = 19) for eligibility assessment by full-text reading. Ultimately, 6 studies were found to be eligible for inclusion in the review (Alladin & Alibhai, 2007; Butler et al., 2008; Chiu et al., 2018; Fuhr et al., 2021; Hernández et al., 2021; Khazraee et al., 2023), according to the criteria established in the registration protocol. An ongoing randomized clinical trial was identified (ACTRN12620000028909), meeting the inclusion criteria; however, reading its registration protocol and contacting the responsible investigator via email confirmed that there are still no results available for inclusion in the review. Another trial was initially listed as probably eligible but was later excluded for being semi-randomized, allowing the patient to decide whether to receive a preferred intervention or be randomly allocated to a group (Dobbin et al., 2009).

**Figure 1.**
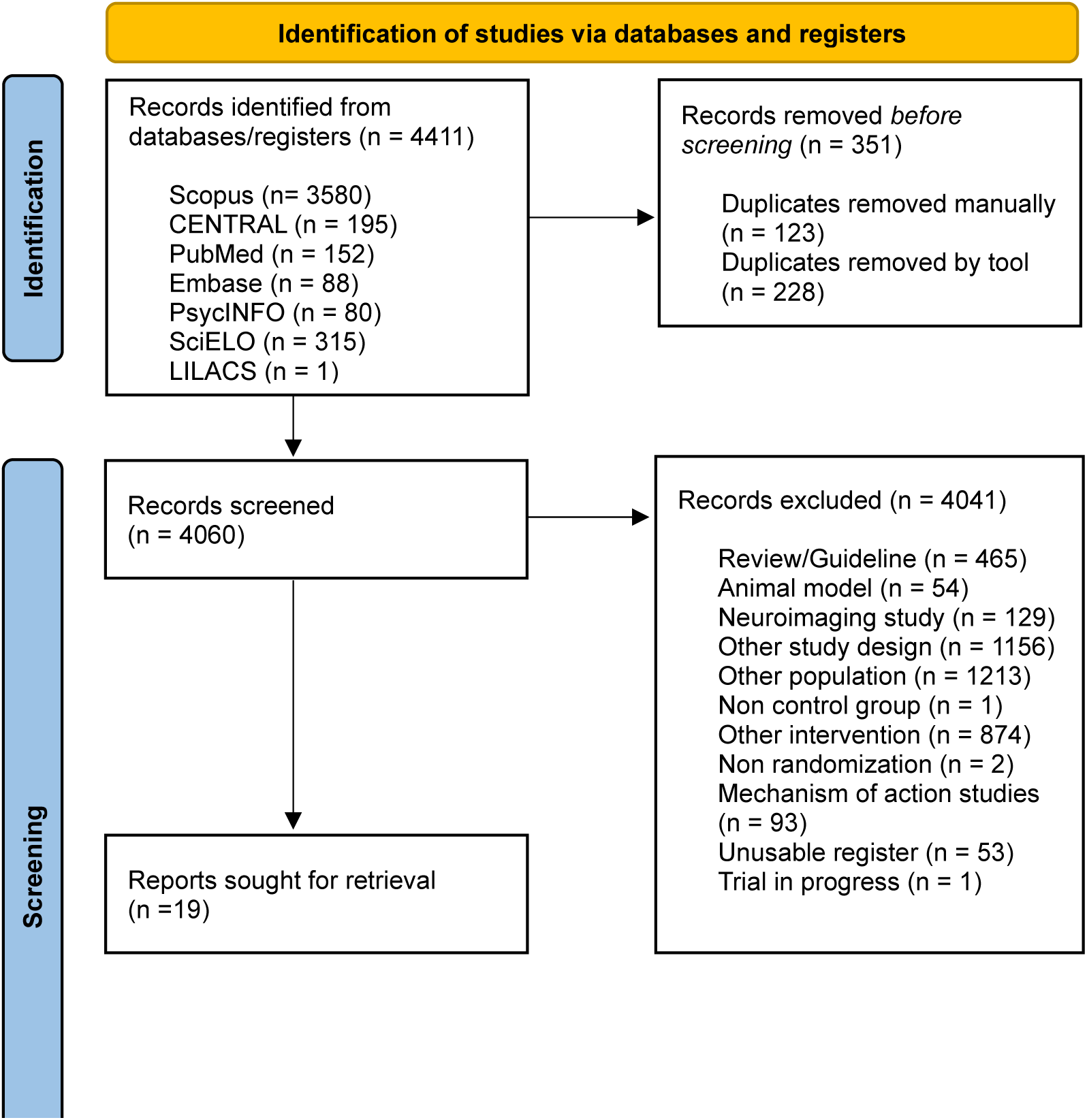
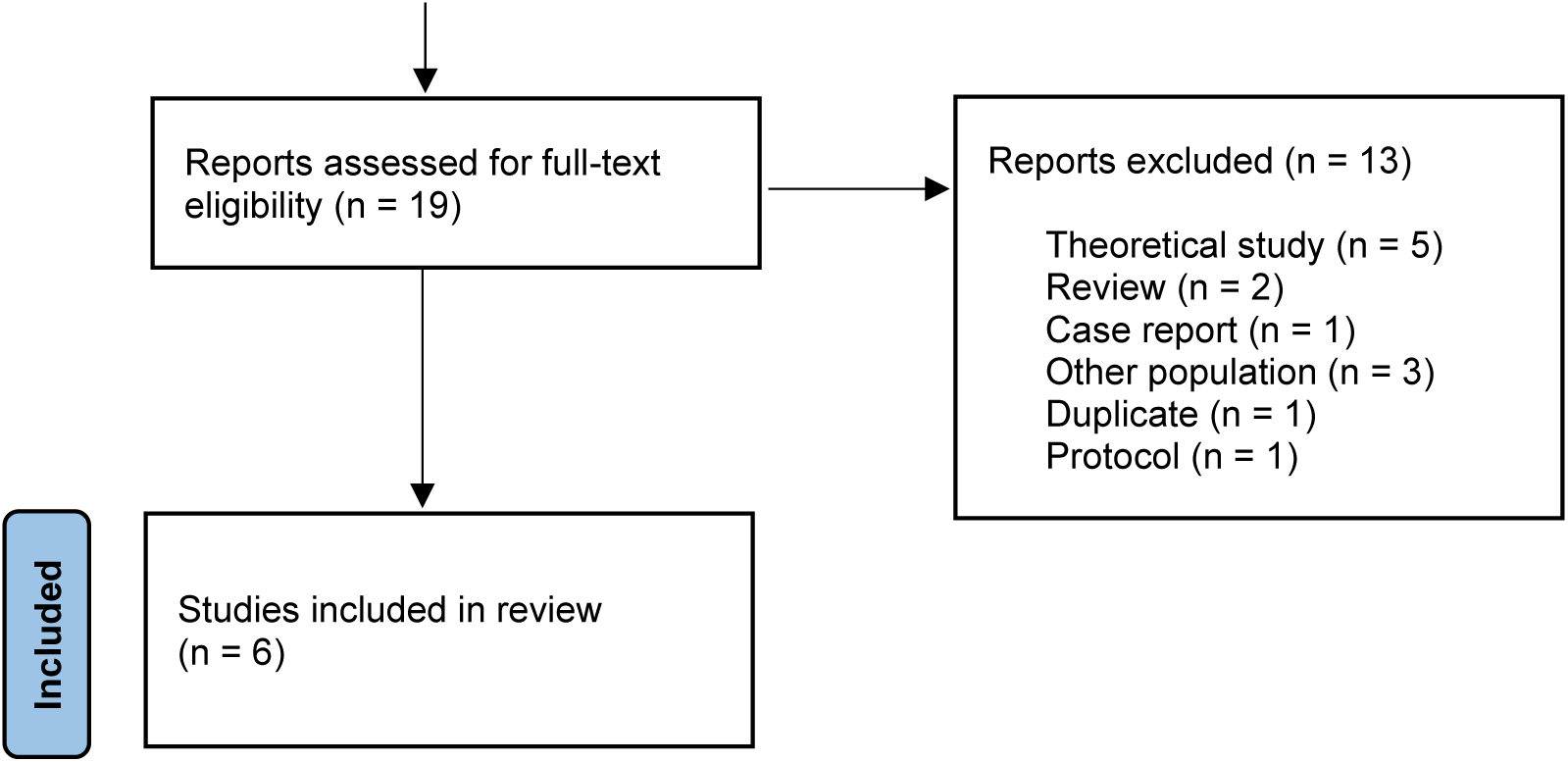
PRISMA 2020 Flow Diagram

### 3.1 DESCRIPTION OF INCLUDED STUDIES

Alladin and Alibhai (2007) conducted an RCT to compare Cognitive Hypnotherapy (CH) to Cognitive Behavioral Therapy in patients with chronic depression (N = 98). HC was superior to CBT in reducing depressive symptoms, anxiety, and hopelessness at the end of the 16-week treatment period, as well as at the 6- and 12-month follow-ups.

Butler et al. (2008) conducted a pilot RCT with two experimental arms compared to a control condition. The intervention for the first experimental group was yoga meditation plus psychoeducation reading materials, the second experimental group was HT plus psychoeducation reading materials, and both interventions were compared to psychoeducation alone in patients (N = 46) with dysthymia (50%), double depression (28%), major depressive episode (15.2%), and chronic depression (6.5%). After 9 months, the yoga group showed significant improvement in remission rates compared to the control group, while the HT group did not show a statistically significant difference. This result may be due to half of the patients in the meditation group and control group receiving treatments external to the experiment and the low sampling of the pilot trial.

Chiu et al. (2018) conducted an RCT comparing HT to pharmacological treatment (Treatment as usual – TAU), the study included 17 (27.4%) patients diagnosed with anxiety, 16 with Mixed Anxiety-Depressive Disorder (25.8%), and finally, 29 (46.8%) of the 62 patients were diagnosed with MDD. The HT group was superior in reducing scores of anxious and depressive symptoms, showing statistically significant differences compared to TAU. However, the authors did not provide a subgroup analysis by diagnosis, preventing an understanding of the therapeutic outcomes that HT achieved in depressed patients without the interference (positive or negative) of results in anxious patients.

Fuhr et al. (2021) conducted a non-inferiority RCT to compare HT to CBT (gold standard treatment) in patients (N = 152) with current episodes of mild to moderate depression, with one patient dropping out of treatment before the first session and being excluded from the analysis. HT was not inferior to CBT at the end of the 20-session treatment in reducing depressive symptoms, maintaining non-inferiority at the 6- and 12-month follow-ups.

Hernández et al. (2021) conducted an RCT to compare HT to Rational Emotive Behavior Therapy (REBT) in depressed patients (N = 30). HT was superior to REBT in remission rates and in reducing symptoms of anxiety, outcomes assessed immediately after the end of the 10 treatment sessions. Khazraee et al. (2023) conducted an RCT to compare Mindful Hypnotherapy (MH), which integrates Mindfulness into HT, versus a waitlist control (WL) in patients (N = 34) diagnosed with depression. MH was superior to WL in reducing depressive symptoms, reducing cognitive inflexibility, and improving self-compassion. This result cannot be generalized to men, as the sample consisted entirely of women.

The six articles, summarized, gather a total sample of N = 422, with N = 214 in the intention-to-treat (ITT) analysis and N = 342 in the per-protocol (PP) analysis with participants who completed the studied treatments. Of the 6 included RCTs, only two (Chiu et al., 2018; Fuhr et al., 2021) presented intention-to-treat analysis. Fuhr et al. also presented per-protocol analysis (hence the simple sum of the N of ITT and the N of PP exceeds N = 422).

### 3.2 INTERVENTIONS

Although four of the included articles could technically be classified as ‘HT’, there is no standardization of therapeutic techniques to be implemented in treatment, and all articles exposed patients to different “settings” for conducting HT, even if it was delivered as a standalone treatment. This review considers all forms of therapeutic intervention involving hypnosis as Hypnosis-Based Interventions (HBI). Alladin and Alibhai (2007) planned a treatment of CBT supplemented by the following hypnotic procedures: hypnotic induction, ego strengthening, expanding awareness, induction of positive mood, post-hypnotic suggestions, and self-hypnosis. The authors did not provide information about participant recruitment or session duration.

Butler et al. (2008) planned a group HT program consisting of weekly sessions for 10 consecutive weeks, each group HT session lasting 1 hour and 30 minutes, with an additional ‘booster’ session of 2 hours in duration in the twelfth week. The sessions were conducted by a psychologist and a psychiatrist, both experienced in hypnosis psychotherapy.

Chiu et al. (2018) designed a HT treatment where patients received 5 to 7 sessions, depending on their clinical condition, over a period of 8 weeks. The scheduled sessions lasted approximately 45 to 75 minutes, during which patients received information about hypnosis and established session goals based on the following dimensions: a) affect; b) cognition; c) behavior; d) biological function; or e) identity. The sessions were conducted using regression and age progression techniques with the application of metaphors and hypnotic suggestions referencing the session goals.

Fuhr et al. (2021) planned a HT treatment delivered in 20 sessions over six months. Unlike CBT, which commonly focuses on rationalizing thoughts, the HT group treatment consisted of emotional activation, reinforcement of personal resources, metaphors, and other techniques described in modules by the authors. Sessions were conducted by professionals with a minimum of 3 years of clinical experience, and treatment fidelity was assessed by four raters to identify whether HT and CBT therapists-maintained adherence to their manuals and used more HT or CBT techniques in their sessions. None of the HT therapists had training in CBT, and vice versa. The study conducted more sessions than are typically administered for patients with mild to moderate depressive symptoms. Normally, for mild depression, 8 to 12 sessions of CBT are delivered, while for moderate cases, 8 to 16 sessions are provided (Gautam et al., 2020). In the study by Fuhr et al. (2021), both the HT and CBT groups received 20 sessions, which is generally indicated for severe depression, a type of patient excluded from the research.

Hernández et al. (2021) designed a HT treatment delivered in 10 sessions, each approximately 45 minutes long, from October 2018 to June 2019. The interventions consisted of applying therapeutic metaphors, suggestions, symbols, and therapeutic resources.

Khazraee et al. (2023) planned a weekly MH treatment consisting of 8 sessions in total, each lasting 60 minutes. The intervention was based on literature referenced by the authors as an MH protocol, with each session’s treatment and suggestions individualized to the patients’ needs and goals. The treatment followed the MH protocol (Elkins & Olendzki, 2018; Olendzki et al., 2020).

### 3.3 COMPARISON

With the exception of Khazraee et al. (2023), which was controlled by a WL, all other studies included in the review were compared to an active control. Two studies (Alladin & Alibhai, 2007; Fuhr et al., 2021) were compared to the gold-standard treatment (CBT), grounded in manuals well-established in the literature (Beck, 1979; Beck, 2002). Butler et al. (2008) compared their two experimental groups to psychoeducation based on a series of internet research about depression and a book on CBT and depression (Burns, 1999); these reading materials were also made available to the experimental patients. The authors Chiu et al. (2018) compared their experimental group to TAU at a psychiatric clinic (antidepressant and anxiolytic medication). Finally, Hernández et al. (2021) compared their experimental group to REBT but did not provide data on the protocol followed or the basis of the control treatment.

### 3.4 OUTCOMES

To assess the outcomes of the treatments, three RCTs (Alladin & Alibhai, 2007; Chiu et al., 2018; Khazraee et al., 2023) used only self-report instruments such as the Beck Depression Inventory (I or II), Beck Anxiety Inventory (BAI), Beck Hopelessness Scale (BHS), and other scales to detect overall improvement effects and anxious symptoms. The RCTs by Butler et al. (2008) and Fuhr et al. (2021) conducted interviews with blind evaluators, using the Hamilton Rating Scale for Depression (HAM-D) and the Montgomery-Åsberg Depression Rating Scale (MADRS), respectively. Hernández et al. (2021) also assessed patients through an interview to evaluate diagnostic status as an outcome, but did not provide further details about the interview.

HT was described as superior in reducing the intensity of depressive symptoms (MD -2.10; 95% CI [-6.16, 1.96]) compared to CBT alone (Alladin & Alibhai, 2007) but the superiority found did not show a significant difference (p = 0.31) between the exposures (see Figure 2). HT supplemented with psychoeducational materials was inferior (see Figure 3) to meditation (MD 1.00; 95% CI [-3.10, 5.10]) also supplemented with psychoeducational materials (Butler et al., 2008), however, besides the difference found not being significant (p = 0.63), the RCT in question is a pilot study, a type of study designed to evaluate the feasibility of researching a particular condition in larger studies that actually have the capacity to find a certain effect (Lee et al., 2014; Kistin & Silverstein, 2015). HT was superior to TAU (MD -8.80; 95% CI [-14.96, -2.64]) prescribed by the psychiatrist of the control group patients (Chiu et al., 2018), being the only study included that achieved a significant difference (p = 0.005) compared to an active treatment group (see Figure 5).

**Figure 2.**
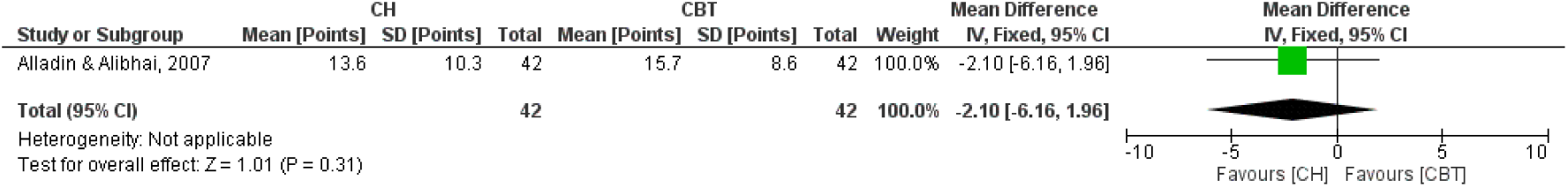
(Analysis 1.1) Descriptive Forest Plot of CH versus CBT.

**Figure 3.**
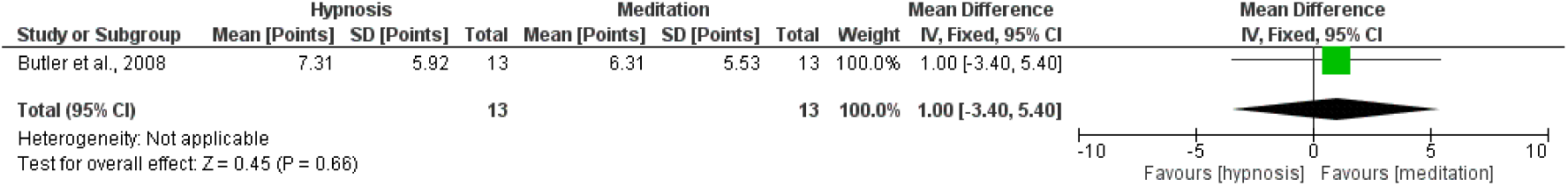
(Analysis 2.1) Descriptive Forest Plot of HT + Psychoeducation versus Yoga Meditation + Psychoeducation.

**Figure 4.**
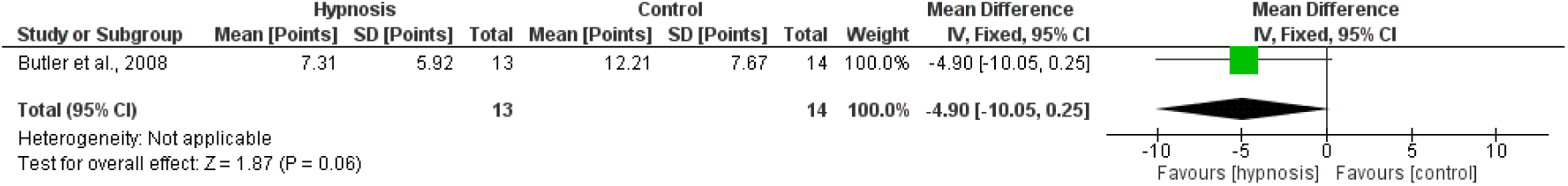
(Analysis 2.2) Descriptive Forest Plot of HT + Psychoeducation versus Psychoeducation Alone.

**Figure 5.**
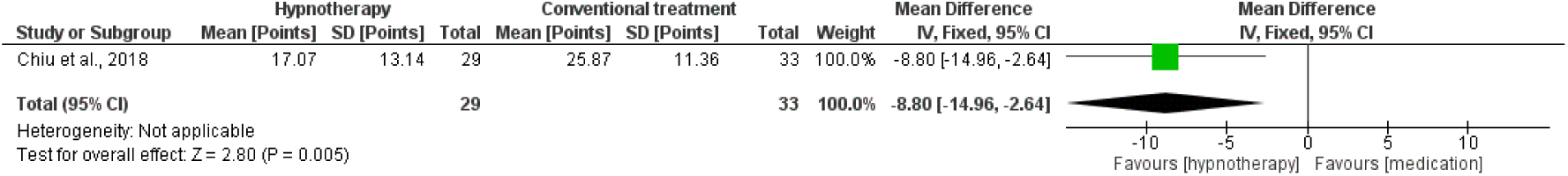
(Analysis 3.1) Descriptive Forest Plot of HT versus TAU (Anxiolytic and Antidepressant Medication).

In the study by Fuhr et al., a non-inferiority RCT was conducted, a type of study that typically aims to compare the effect of a new treatment to the gold standard to evaluate the non-inferiority or superiority of the new treatment, potentially justifying the prescription of an intervention over the established treatment for reasons beyond efficacy, such as accessibility to ‘treatment B’ (Kim et al., 2022). The non-inferiority margin was set at ½ SD (Fuhr et al., 2017), resulting in a slightly better but not significant outcome (p = 0.95) in favor of HT, confirming its non-inferiority (see Figure 6) compared to CBT (MD -0.10; 95% CI [-2.99, 2.79]). The authors of the trial also conducted an evaluation of the functional changes in brain activity of participants before and after the intervention through near-infrared functional spectroscopy, demonstrating that the HT and CBT groups had clinically non-significant different results but with different changes in functional connectivity depending on the magnitude of mental rumination. HT altered the activity of the Superior Temporal Sulcus (STS), while CBT helped normalize prefrontal and amygdala activity (Haipt et al., 2022). For a better understanding of the possible therapy outcomes, the authors extended the follow-up in a survival analysis over 12 months with 136 patients available for analysis, where HT was comparable to CBT in maintaining low levels of depressive symptoms and achieving high long-term remission rates, with both treatments resulting in 73% remission after a median of 30 weeks (Fuhr et al., 2023).

**Figure 6.**
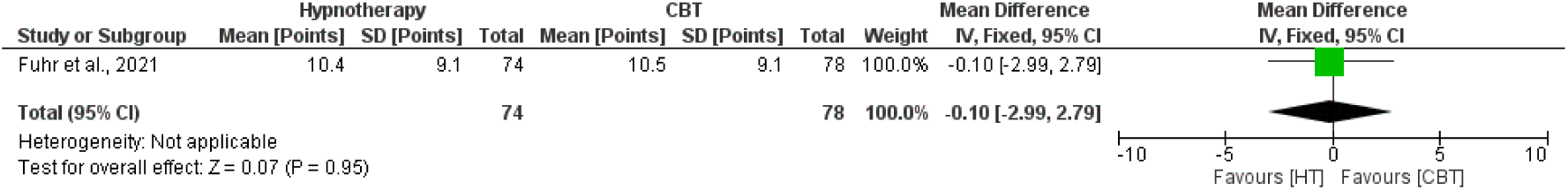
(Analysis 4.1) Descriptive Forest Plot of HT versus CBT in a Non-Inferiority Clinical Trial in ITT Analysis.

**Figure 7.**
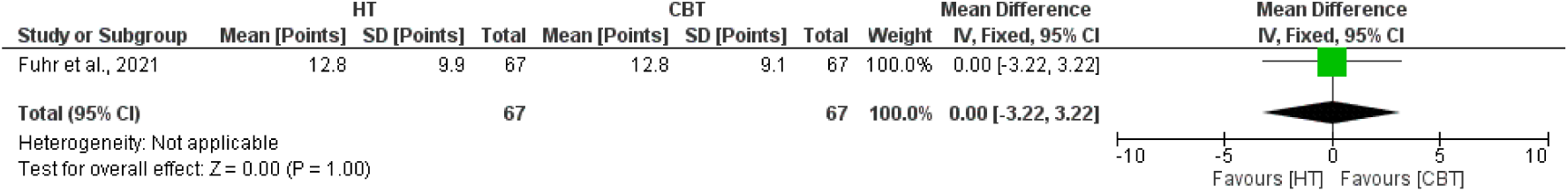
(Analysis 4.2) Descriptive Forest Plot of HT versus CBT in a Non-Inferiority Clinical Trial in PP Analysis.

In the study by Hernández et al., a categorical outcome analysis revealed that HT was associated with an 80% protective effect in reducing the odds of no improvement compared to REBT (OR 0.20; 95% CI [0.02, 2.02]). Although the adjusted odds ratio suggests a favorable trend towards the experimental treatment, this difference did not reach statistical significance (p = 0.17), indicating that there is not enough evidence to claim a clear superiority of the experimental treatment over the control based on this study (see Figure 8). The study by Khazraee et al. was the only RCT included in the review that had its experimental group compared to a non-active group, demonstrating a large effect size (MD -25.56, CI 95% [-30.78, -20.34]) compared to the control (see Figure 9), achieving a significant difference.

**Figure 8.**
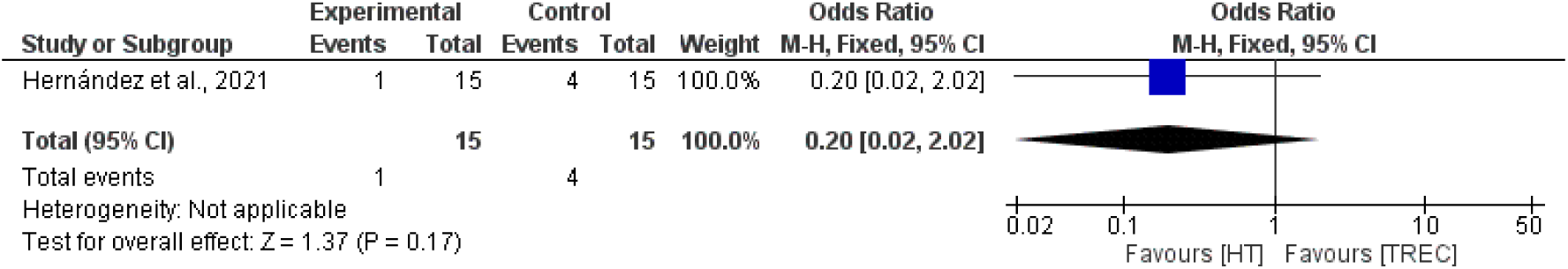
(Analysis 5.1) Descriptive Forest Plot of HT versus REBT. Categorical Outcome: Remaining Diagnosed with MDD or Not at Follow-Up.

**Figure 9.**
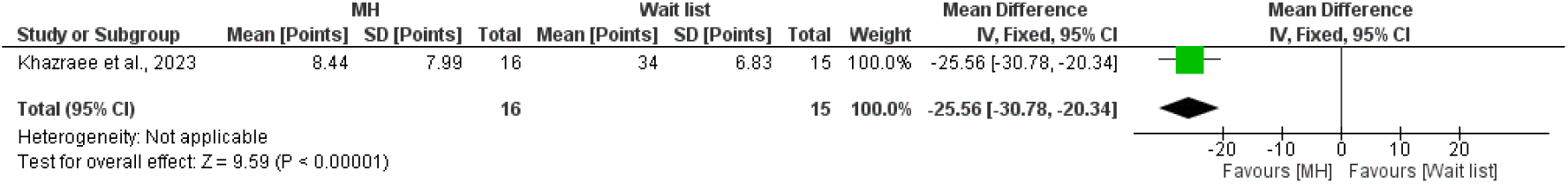
(Analysis 6.1) Descriptive Forest Plot of MH versus Waitlist.

### 3.5 RISK OF BIAS

The included articles were assessed by two independent investigators (F.L.S. and M.S.M.) using the RoB 2 tool (<u>Cochrane Methods</u>) after receiving training from two senior researchers with experience in publishing Cochrane systematic reviews. The overall assessment of the evidence included in the review was judged to have a high risk of bias (see Figure 12).

**Figure 11.**
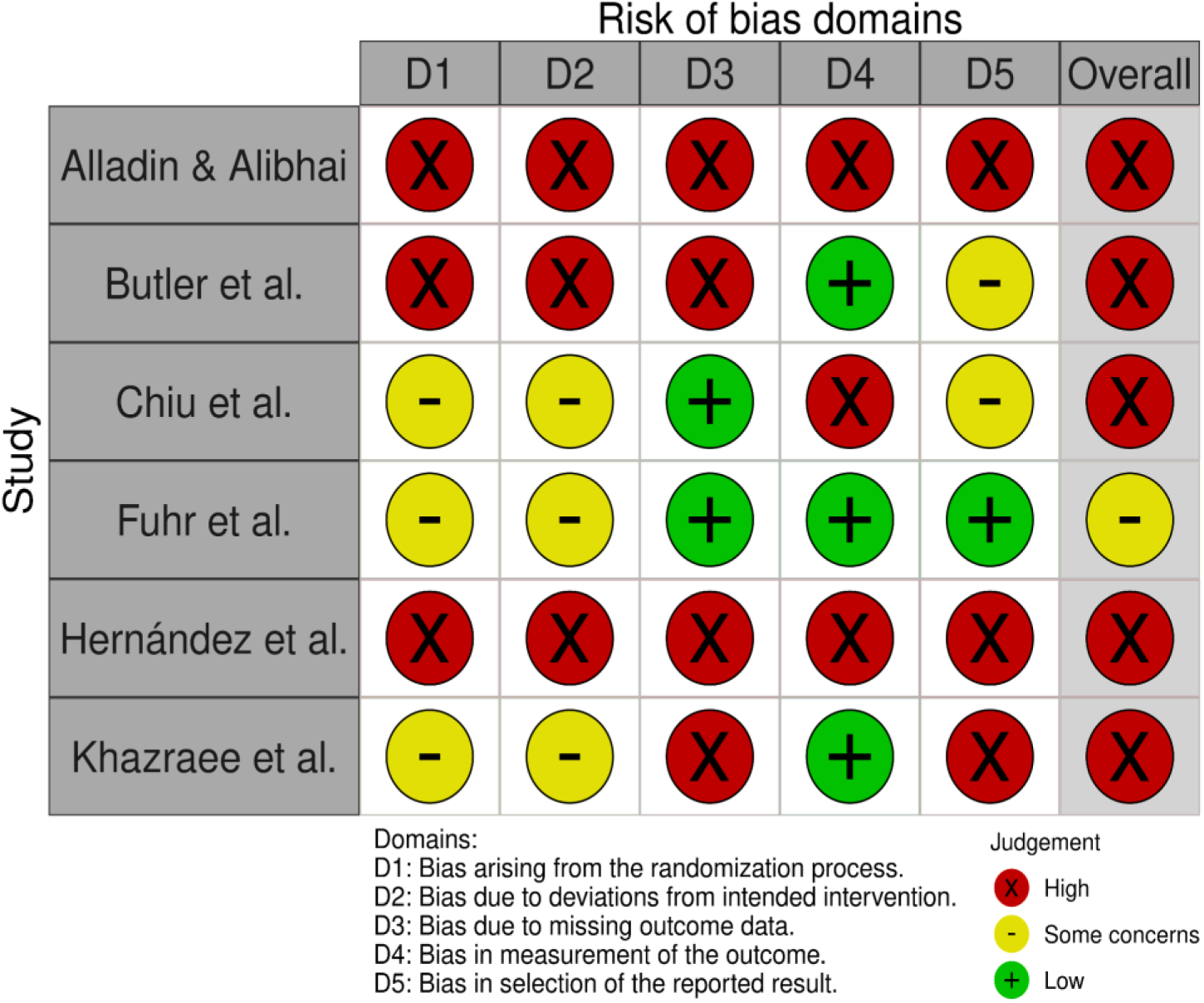
RoB 2 Traffic Light Plot for Risk of Bias Assessment.

**Figure 12.**
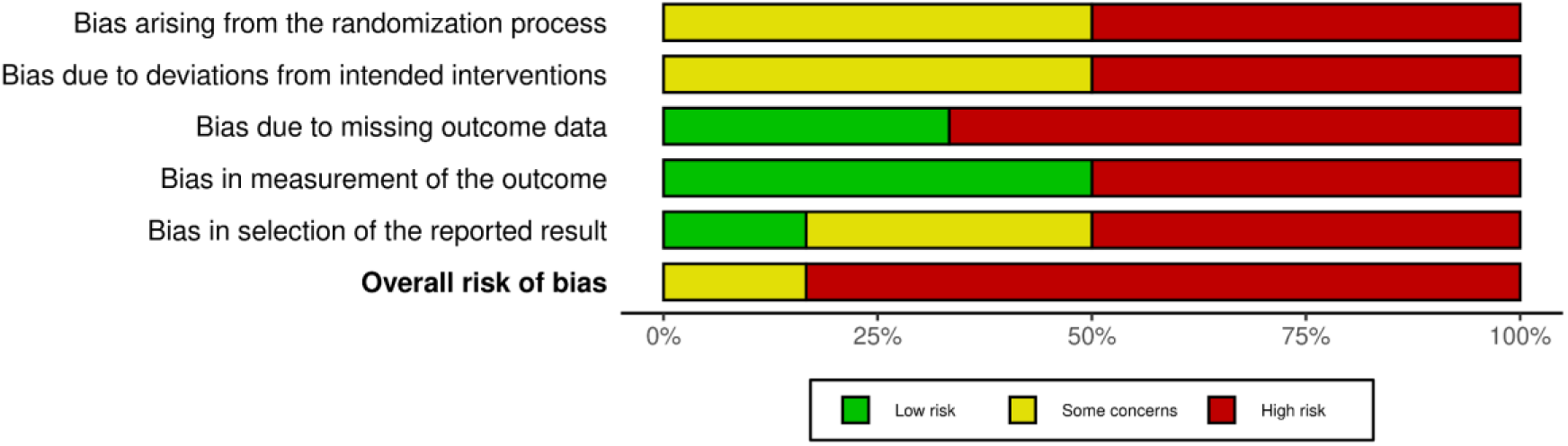
Summary Plot of the Risk of Bias for Included Randomized Clinical Trials.

[D1] Of the reviewed studies, only the RCT by Fuhr et al. provided sufficient information for an adequate assessment of the randomization process domain, where an external biometrics institute generated the randomization of participants. The studies by Alladin & Alibhai, Hernández et al., and Chiu et al. stated that their patients were randomly allocated to their groups (intervention or control) but did not provide details on the randomization and allocation process. Butler et al. described the randomization process but stated that the allocation was not concealed. Khazraee et al. did not provide satisfactory details about the sociodemographic data between groups to assess the randomization.

[D2] The study by Fuhr et al. presents evidence of significant prognostic imbalance (such as diagnostic subtypes, medications, and others) between the groups, raising concerns about the presence of confounding factors that could influence the study outcome. This creates doubts about a breakdown in the randomization process and a deviation from the intended intervention. Additionally, the asymmetry in medication use during the study was not balanced between the groups. In Chiu et al., the prognostic imbalance (similar to that in Fuhr et al., 2021) also raises concerns about deviation from the intended interventions due to confounding factors, and the sociodemographic data were presented without their standard deviations. All other studies do not provide adequate data for assessment.

[D3] Only two studies conducted an ITT analysis (Chiu et al., 2018), with one of these performing both ITT and PP analyses and satisfactorily clarifying the missing data and how they were imputed for analysis (Fuhr et al., 2021). The other studies conducted PP analysis without providing adequate information for evaluation.

[D4] A pre-registration of the analysis plan was identified only for the study by Fuhr et al. (Fuhr et al., 2017). Three studies used only self-report measures (Alladin & Alibhai, 2007; Chiu et al., 2018; Khazraee et al., 2023), resulting in a high risk of bias assessment. However, the Iranian study (Khazraee et al., 2023) provided sufficient data on the pre- and post-intervention data collection by assessors blinded to the study’s objective, establishing a low risk of bias assessment, unlike the other self-report studies which did not provide adequate information on data collection and/or used outdated instruments compared to their more recent versions (available at the time of the study). The study by Fuhr et al. and the study by Butler et al. used evaluations based on observers blinded to the study conditions. The RCT by Hernández et al. also used evaluations based on clinical observers but provided insufficient information for process evaluation; additionally, it applied a self-report scale to assess anxiety intensity.

[D5] The mere absence of an analysis plan would classify the studies as having “some concerns” in the risk of bias assessment. However, certain details exacerbated the risk of bias judgment. The study by Alladin & Alibhai does not even report the number of participants in the groups throughout the follow-up time points, nor does it explain how these data were collected. Khazraee et al. (2023) report only the results of three out of the nine outcomes listed as primary outcomes in the Iranian Registry of Clinical Trials (IRCT20211210053342N1), which highlights the use of nine psychometric tools (including scales, questionnaires, and inventories). Fuhr et al. provided all the pre-specified data in the analysis plan (Fuhr et al., 2017).

### 3.6 CERTAINTY OF EVIDENCE

As planned (CRD42023409631), this review used the GRADE approach to assess the quality of evidence and judge the recommendation of hypnosis-based interventions for depression, resulting in 7 different PICOTS.

**Table 2.**
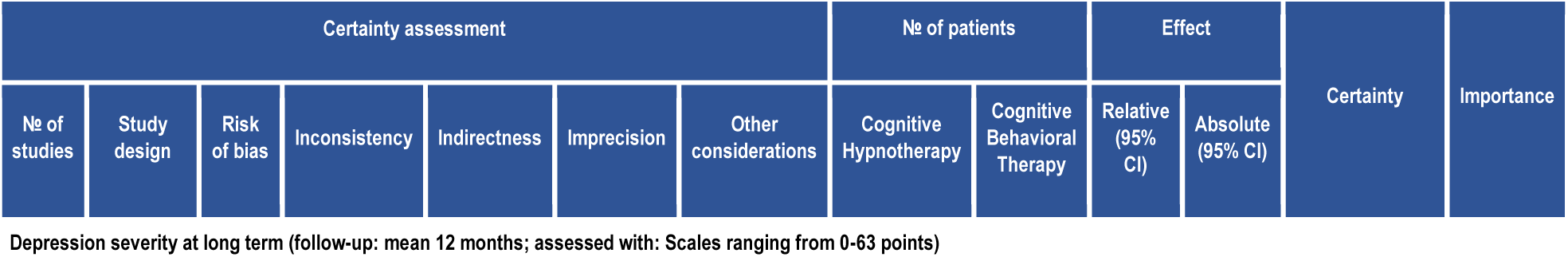
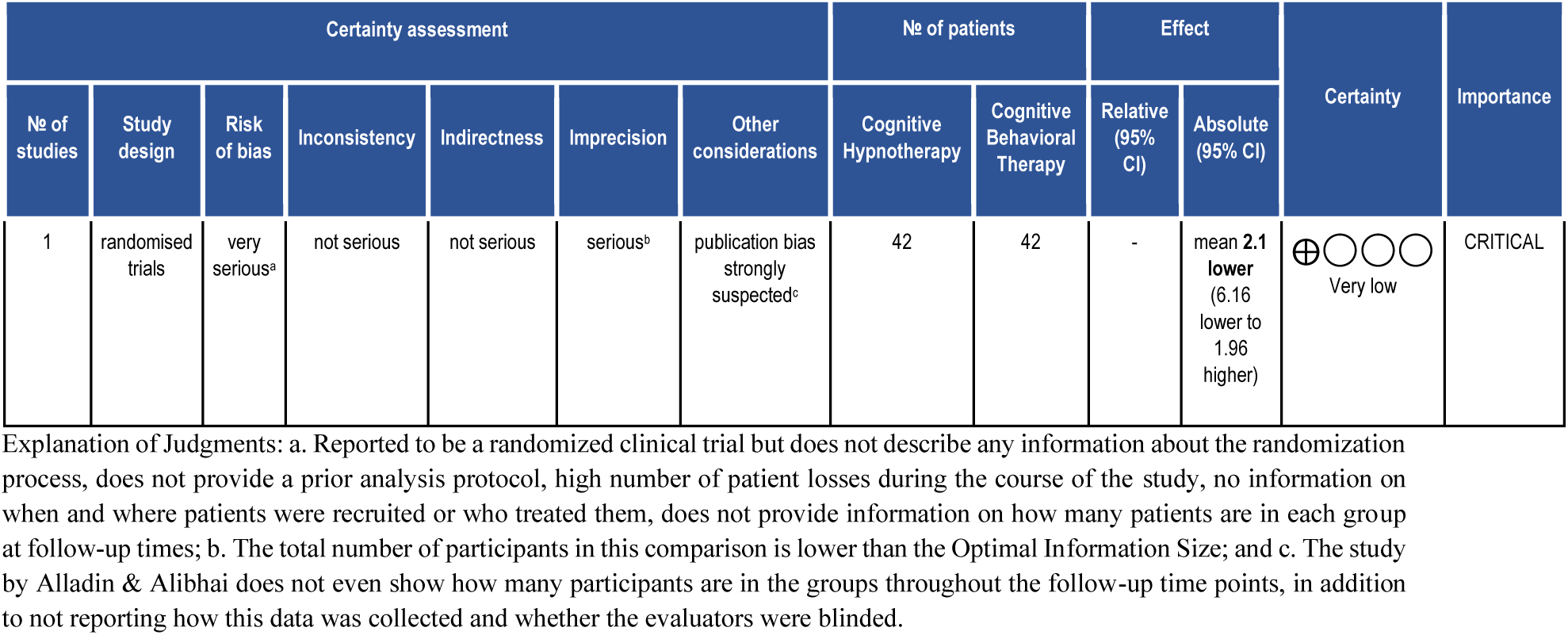
GRADE for CH versus CBT.

| Certainty assessment |  |  |  |  |  |  | No of patients |  | Effect |  | Certainty | Importance |
| --- | --- | --- | --- | --- | --- | --- | --- | --- | --- | --- | --- | --- |
| No of studies | Study design | Risk of bias | Inconsistency | Indirectness | Imprecision | Other considerations | Cognitive Hypnotherapy | Cognitive Behavioral Therapy | Relative (95% CI) | Absolute (95% CI) |  |  |

Depression severity at long term (follow-up: mean 12 months; assessed with: Scales ranging from 0-63 points)
| Certainty assessment |  |  |  |  |  |  | № of patients |  | Effect |  | Certainty | Importance |
| --- | --- | --- | --- | --- | --- | --- | --- | --- | --- | --- | --- | --- |
| № of studies | Study design | Risk of bias | Inconsistency | Indirectness | Imprecision | Other considerations | Cognitive Hypnotherapy | Cognitive Behavioral Therapy | Relative (95% CI) | Absolute (95% CI) |  |  |
| 1 | randomised trials | very serious<sup>a</sup> | not serious | not serious | serious<sup>b</sup> | publication bias strongly suspected<sup>c</sup> | 42 | 42 | - | mean 2.1 lower (6.16 lower to 1.96 higher) | ⊕○○○<br>Very low | CRITICAL |

Explanation of Judgments: a. Reported to be a randomized clinical trial but does not describe any information about the randomization process, does not provide a prior analysis protocol, high number of patient losses during the course of the study, no information on when and where patients were recruited or who treated them, does not provide information on how many patients are in each group at follow-up times; b. The total number of participants in this comparison is lower than the Optimal Information Size; and c. The study by Alladin & Alibhai does not even show how many participants are in the groups throughout the follow-up time points, in addition to not reporting how this data was collected and whether the evaluators were blinded.

**Table 3.**
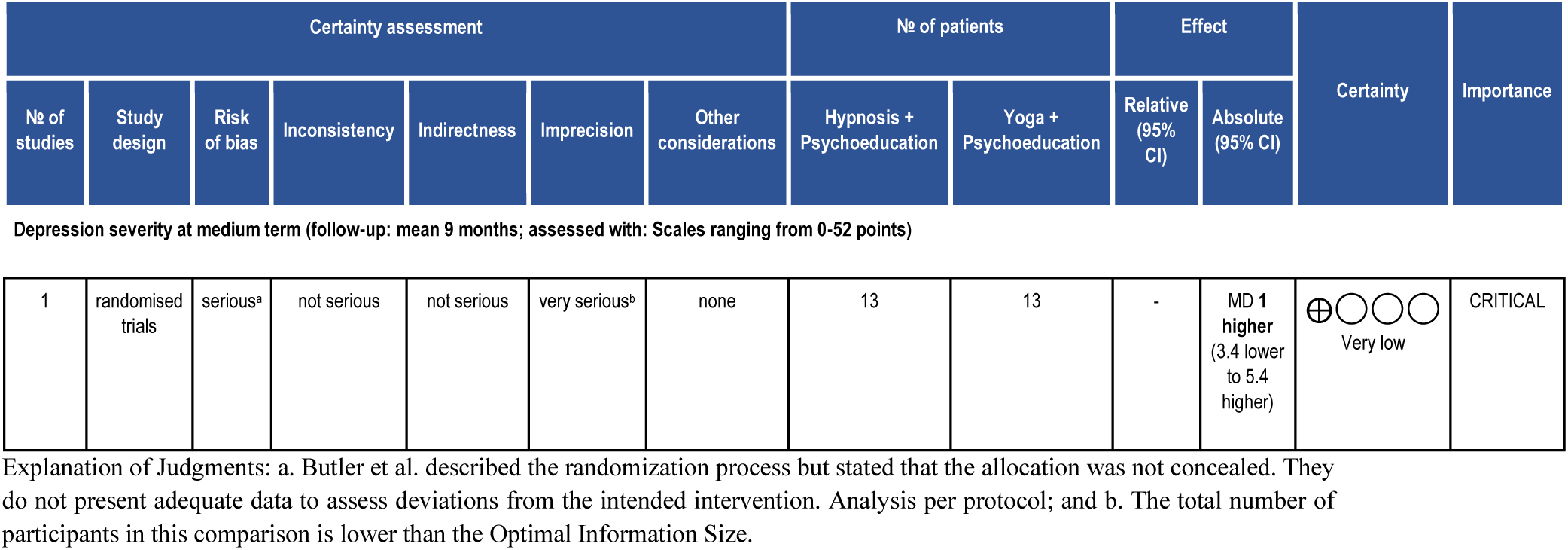
GRADE for HT + Psychoeducation versus Yoga Meditation + Psychoeducation.

| Certainty assessment |  |  |  |  |  |  | № of patients |  | Effect |  | Certainty | Importance |
| --- | --- | --- | --- | --- | --- | --- | --- | --- | --- | --- | --- | --- |
| № of studies | Study design | Risk of bias | Inconsistency | Indirectness | Imprecision | Other considerations | Hypnosis + Psychoeducation | Yoga + Psychoeducation | Relative (95% CI) | Absolute (95% CI) |  |  |
| Depression severity at medium term (follow-up: mean 9 months; assessed with: Scales ranging from 0-52 points) |  |  |  |  |  |  |  |  |  |  |  |  |
| 1 | randomised trials | serious<sup>a</sup> | not serious | not serious | very serious<sup>b</sup> | none | 13 | 13 | - | MD 1 higher (3.4 lower to 5.4 higher) | ⊕○○○<br>Very low | CRITICAL |

Explanation of Judgments: a. Butler et al. described the randomization process but stated that the allocation was not concealed. They do not present adequate data to assess deviations from the intended intervention. Analysis per protocol; and b. The total number of participants in this comparison is lower than the Optimal Information Size.

**Table 4.**
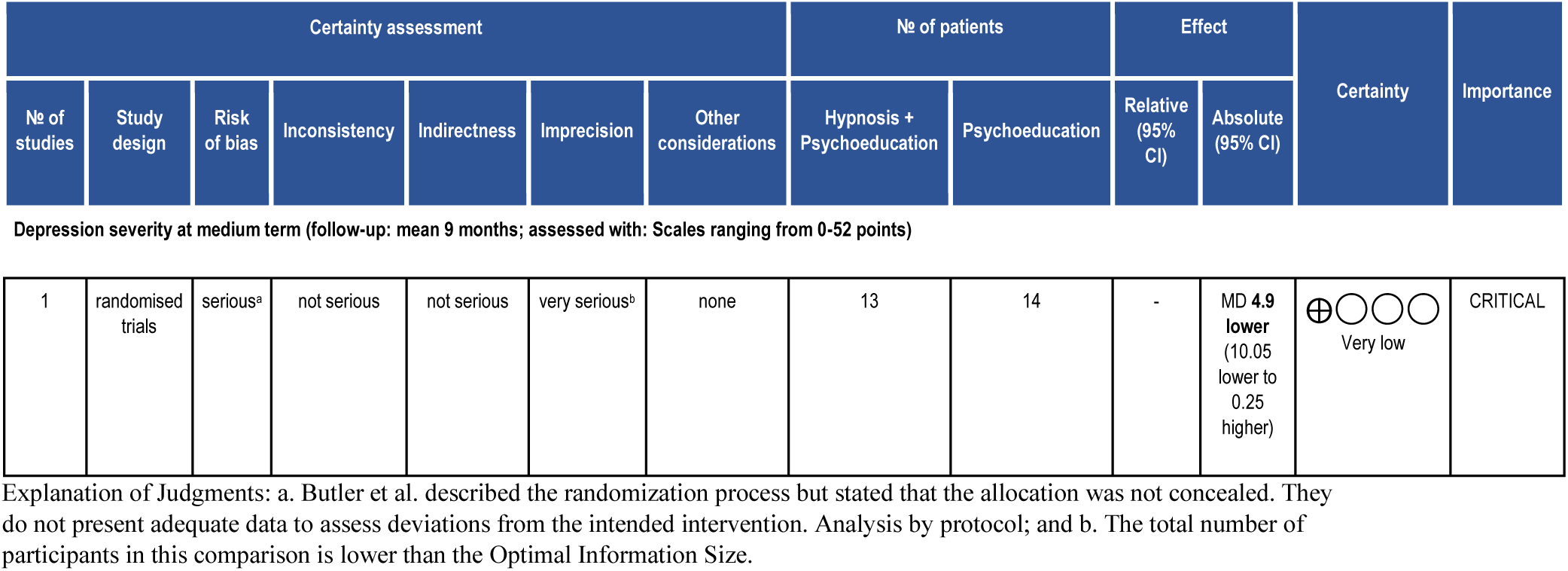
GRADE for HT + Psychoeducation versus Psychoeducation.

| Certainty assessment |  |  |  |  |  |  | № of patients |  | Effect |  | Certainty | Importance |
| --- | --- | --- | --- | --- | --- | --- | --- | --- | --- | --- | --- | --- |
| № of studies | Study design | Risk of bias | Inconsistency | Indirectness | Imprecision | Other considerations | Hypnosis + Psychoeducation | Psychoeducation | Relative (95% CI) | Absolute (95% CI) |  |  |
| Depression severity at medium term (follow-up: mean 9 months; assessed with: Scales ranging from 0-52 points) |  |  |  |  |  |  |  |  |  |  |  |  |
| 1 | randomised trials | serious<sup>a</sup> | not serious | not serious | very serious<sup>b</sup> | none | 13 | 14 | - | MD 4.9 lower (10.05 lower to 0.25 higher) | ⊕○○○<br>Very low | CRITICAL |

Explanation of Judgments: a. Butler et al. described the randomization process but stated that the allocation was not concealed. They do not present adequate data to assess deviations from the intended intervention. Analysis by protocol; and b. The total number of participants in this comparison is lower than the Optimal Information Size.

**Table 5.**
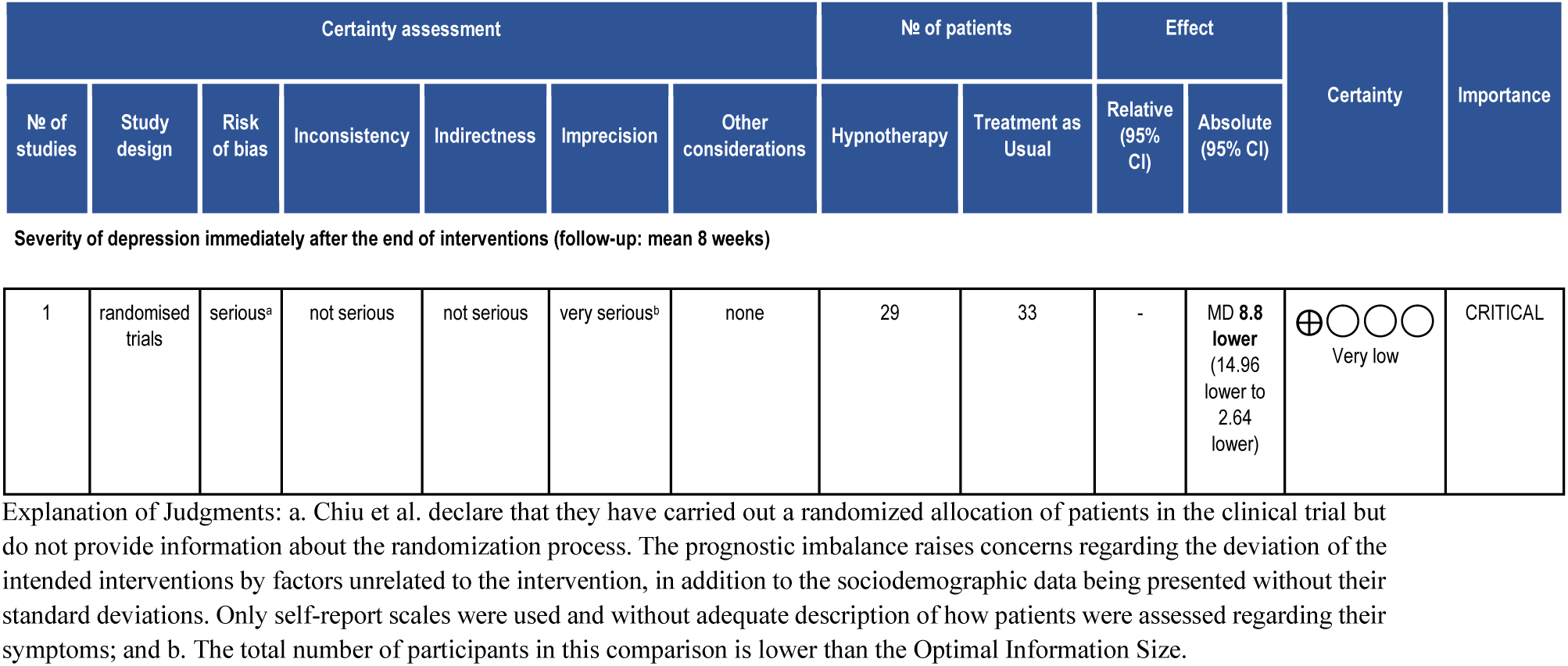
GRADE for HT versus TAU.

| Certainty assessment |  |  |  |  |  |  | № of patients |  | Effect |  | Certainty | Importance |
| --- | --- | --- | --- | --- | --- | --- | --- | --- | --- | --- | --- | --- |
| № of studies | Study design | Risk of bias | Inconsistency | Indirectness | Imprecision | Other considerations | Hypnotherapy | Treatment as Usual | Relative (95% CI) | Absolute (95% CI) |  |  |
| Severity of depression immediately after the end of interventions (follow-up: mean 8 weeks) |  |  |  |  |  |  |  |  |  |  |  |  |
| 1 | randomised trials | serious<sup>a</sup> | not serious | not serious | very serious<sup>b</sup> | none | 29 | 33 | - | MD 8.8 lower (14.96 lower to 2.64 lower) | ⊕○○○<br>Very low | CRITICAL |

Explanation of Judgments: a. Chiu et al. declare that they have carried out a randomized allocation of patients in the clinical trial but do not provide information about the randomization process. The prognostic imbalance raises concerns regarding the deviation of the intended interventions by factors unrelated to the intervention, in addition to the sociodemographic data being presented without their standard deviations. Only self-report scales were used and without adequate description of how patients were assessed regarding their symptoms; and b. The total number of participants in this comparison is lower than the Optimal Information Size.

**Table 6.**
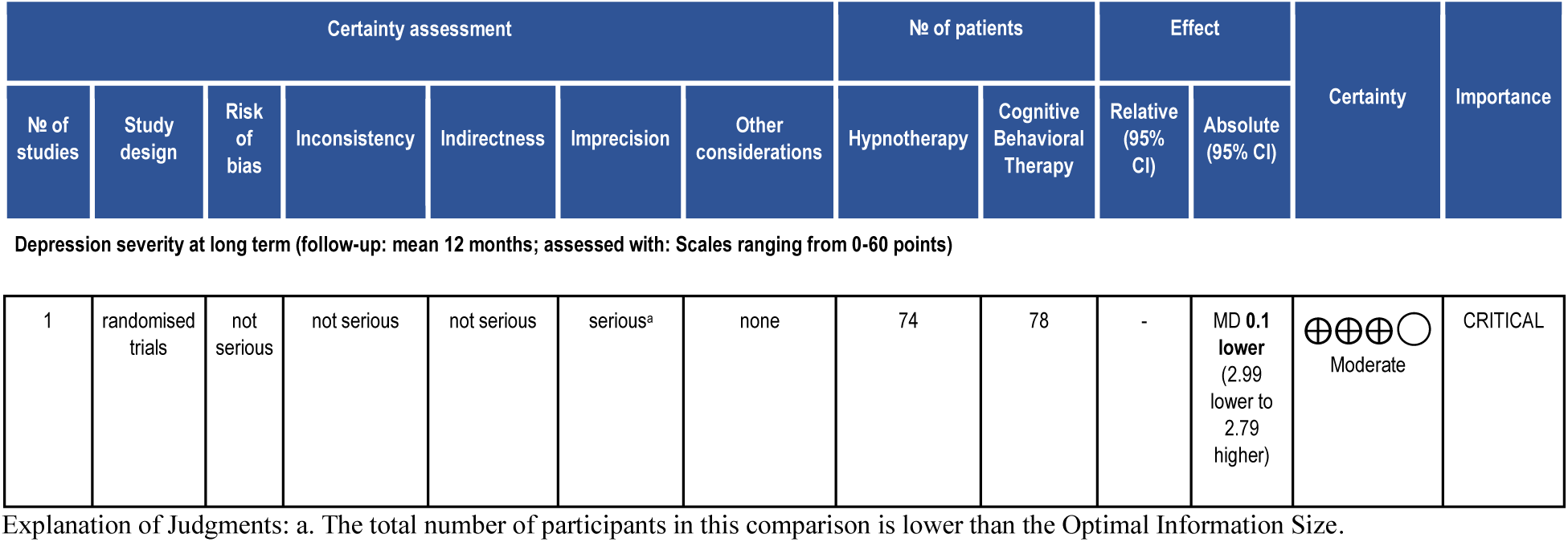
GRADE for HT versus CBT.

| Certainty assessment |  |  |  |  |  |  | № of patients |  | Effect |  | Certainty | Importance |
| --- | --- | --- | --- | --- | --- | --- | --- | --- | --- | --- | --- | --- |
| № of studies | Study design | Risk of bias | Inconsistency | Indirectness | Imprecision | Other considerations | Hypnotherapy | Cognitive Behavioral Therapy | Relative (95% CI) | Absolute (95% CI) |  |  |
| Depression severity at long term (follow-up: mean 12 months; assessed with: Scales ranging from 0-60 points) |  |  |  |  |  |  |  |  |  |  |  |  |
| 1 | randomised trials | not serious | not serious | not serious | serious<sup>a</sup> | none | 74 | 78 | - | MD 0.1 lower (2.99 lower to 2.79 higher) | ⊕⊕⊕○<br>Moderate | CRITICAL |

Explanation of Judgments: a. The total number of participants in this comparison is lower than the Optimal Information Size.

**Table 7.**
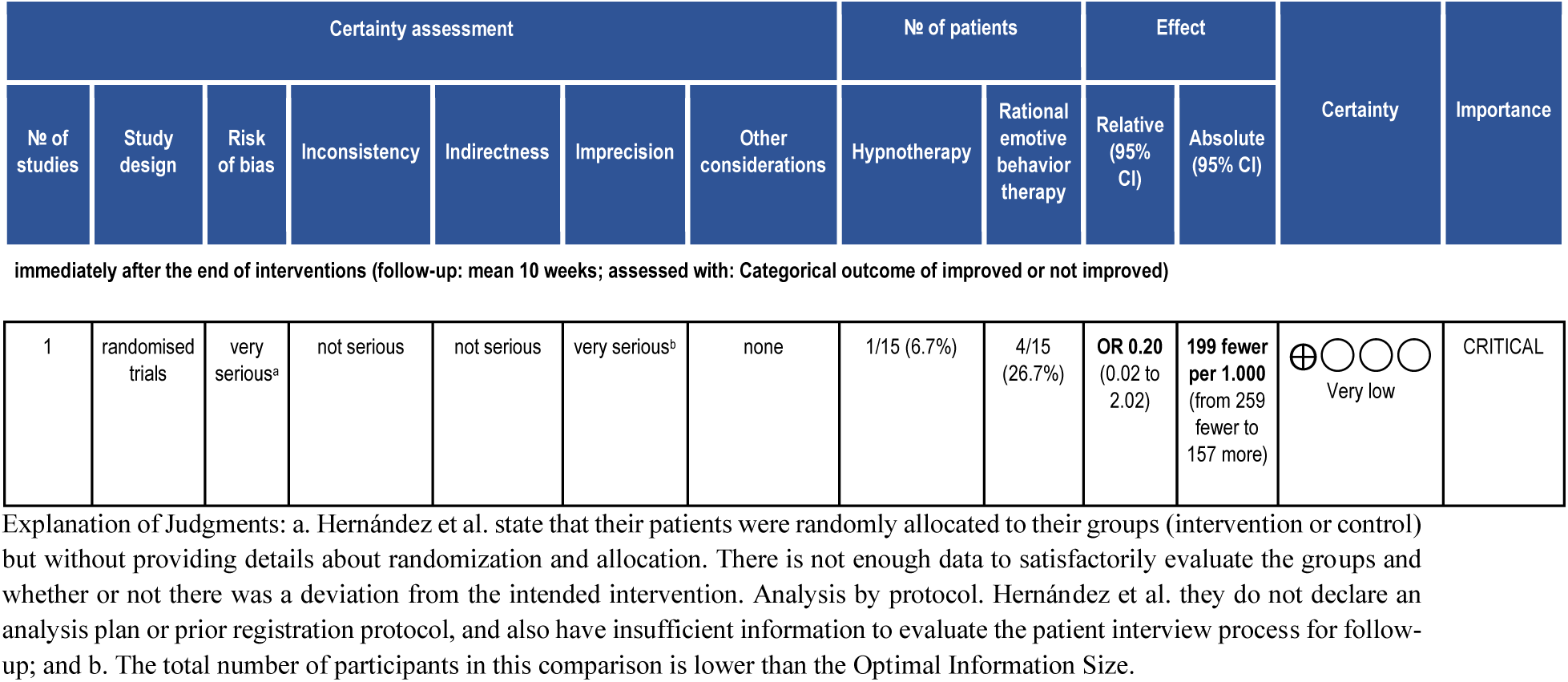
GRADE for HT versus REBT.

| Certainty assessment |  |  |  |  |  |  | № of patients |  | Effect |  | Certainty | Importance |
| --- | --- | --- | --- | --- | --- | --- | --- | --- | --- | --- | --- | --- |
| № of studies | Study design | Risk of bias | Inconsistency | Indirectness | Imprecision | Other considerations | Hypnotherapy | Rational emotive behavior therapy | Relative (95% CI) | Absolute (95% CI) |  |  |
| 1 | randomised trials | very serious<sup>a</sup> | not serious | not serious | very serious<sup>b</sup> | none | 1/15 (6.7%) | 4/15 (26.7%) | OR 0.20 (0.02 to 2.02) | 199 fewer per 1,000 (from 259 fewer to 157 more) | ⊕○○○<br>Very low | CRITICAL |

Explanation of Judgments: a. Hernández et al. state that their patients were randomly allocated to their groups (intervention or control) but without providing details about randomization and allocation. There is not enough data to satisfactorily evaluate the groups and whether or not there was a deviation from the intended intervention. Analysis by protocol. Hernández et al. they do not declare an analysis plan or prior registration protocol, and also have insufficient information to evaluate the patient interview process for follow-up; and b. The total number of participants in this comparison is lower than the Optimal Information Size.

**Table 8.**
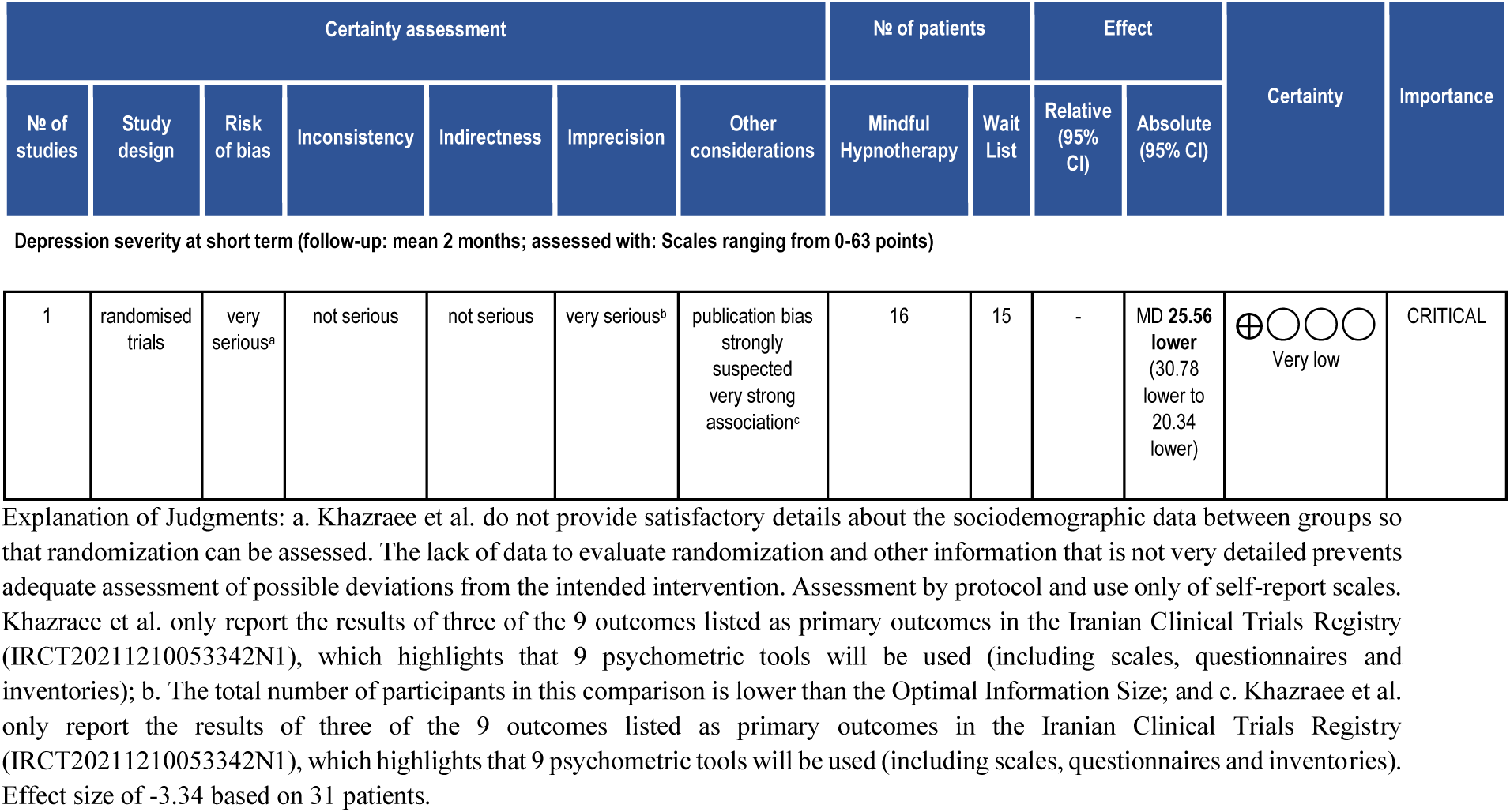
GRADE for MH versus WL.

| Certainty assessment |  |  |  |  |  |  | № of patients |  | Effect |  | Certainty | Importance |
| --- | --- | --- | --- | --- | --- | --- | --- | --- | --- | --- | --- | --- |
| № of studies | Study design | Risk of bias | Inconsistency | Indirectness | Imprecision | Other considerations | Mindful Hypnotherapy | Wait List | Relative (95% CI) | Absolute (95% CI) |  |  |
| Depression severity at short term (follow-up: mean 2 months; assessed with: Scales ranging from 0-63 points) |  |  |  |  |  |  |  |  |  |  |  |  |
| 1 | randomised trials | very serious<sup>a</sup> | not serious | not serious | very serious<sup>b</sup> | publication bias strongly suspected very strong association<sup>c</sup> | 16 | 15 | - | MD 25.56 lower (30.78 lower to 20.34 lower) | Very low | CRITICAL |

Explanation of Judgments: a. Khazraee et al. do not provide satisfactory details about the sociodemographic data between groups so that randomization can be assessed. The lack of data to evaluate randomization and other information that is not very detailed prevents adequate assessment of possible deviations from the intended intervention. Assessment by protocol and use only of self-report scales. Khazraee et al. only report the results of three of the 9 outcomes listed as primary outcomes in the Iranian Clinical Trials Registry (IRCT20211210053342N1), which highlights that 9 psychometric tools will be used (including scales, questionnaires and inventories); b. The total number of participants in this comparison is lower than the Optimal Information Size; and c. Khazraee et al. only report the results of three of the 9 outcomes listed as primary outcomes in the Iranian Clinical Trials Registry (IRCT20211210053342N1), which highlights that 9 psychometric tools will be used (including scales, questionnaires and inventories). Effect size of -3.34 based on 31 patients.

## 4. DISCUSSION

The included studies evaluated the effect of HT (with significant heterogeneity in its formats) on reducing the intensity of depressive symptoms and changing diagnostic status (remission). In general, HC did not show statistically significant differences compared to CBT, and HT did not show statistically significant differences compared to meditation or psychoeducation, nor did it show differences compared to REBT. HT demonstrated statistically significant differences compared to TAU and waitlist control, supposedly indicating its superiority over these controls. However, these results are derived from studies with a high risk of bias, making the credibility of the results questionable. The non-inferiority of HT compared to CBT was confirmed, but this result should be interpreted with caution due to methodological concerns.

The studies also assessed secondary outcomes not discussed in this work as they fall outside the scope of the targeted response, avoiding the perception of “spin” in this review by readers (Siex et al., 2020; Yavchitz et al., 2016). This review included only RCTs as they are the gold standard for evaluating the efficacy and effectiveness of health interventions (Schulz et al., 2010; Djulbegovic & Guyatt, 2017).

Evidence-based practices exert a strong clinical, educational, and experimental influence by providing ethical and empirically supported treatments. Although similar, these practices (EBPs and ESTs) are distinct concepts, and many professionals struggle to differentiate them in their practice (Drisko & Friedman, 2019). Despite empirically supported treatments (ESTs) proving effective outside of RCTs, the mere production of more evidence for ESTs is insufficient for their community application, as real-world therapist confidence in ESTs is limited outside academia, reducing their practical adoption (Schneider et al., 2020).

The criteria for endorsing an intervention as empirically supported by the Division 12 of the American Psychological Association (APA) are broad, provoking numerous criticisms and discussions. An intervention is considered an EST if it: 1) has two or more RCTs conducted by different research teams demonstrating superiority to placebo, another psychotherapeutic intervention, or equivalence to an established treatment with statistical significance; or 2) has nine or more single-case studies conducted by different researchers showing superiority to placebo or other established treatments (Chambless & Hollon, 1998). In this context, hypnotherapy has not gathered randomized evidence over the past two decades to be classified as an EST for the treatment of depression.

The poor methodological quality of the RCTs undermines the credibility of their results, whether due to the planning of data collection, the conduct of the research, or the reporting of results. Even serving as possible proponents for new research to verify their results with better designs, these studies are inadequate for providing credible scientific evidence. Such research is common in biomedical literature, characterized by the perpetuation of non-applicable, inconclusive, and invalid results - in other words, false (Ioannidis, 2005).

Two previously published meta-analyses indicated HT as a potentially effective treatment for depressive symptoms (Shih et al., 2009; Milling et al., 2019), while a third suggested HC as superior to CBT alone (Ramondo et al., 2021). However, their results stem from comparisons of studies with substantial heterogeneity (e.g., Shih et al. included trials with depressed versus non-depressed groups, cancer patients, depressed students, etc.), excessively lenient judgments regarding the risk of bias in the studies (e.g., Ramondo et al. categorized studies that provided no information on the randomization process as having “some concerns”), and extrapolated claims about the quality of the incorporated evidence (e.g., Milling et al. concluded that HT is a very effective intervention based on underpowered studies with high risk of bias). These meta-analyses also included dissertations and studies conducted with patients without a formal diagnosis of depressive disorder.

This review categorically states that based on the available evidence, the potential effect of HT (alone or supplemented) in treating depression is unknown, and new studies are very likely to change our certainty in the estimates of HT effects. Despite this scenario, the German trial of HT versus CBT (Fuhr et al., 2021) stands out from the other trials included in the review not only due to the weight of its sample size but also because of the publication of a protocol with an analysis plan and a detailed description of the interventions. It was classified as having “some concerns” regarding its bias due to the previously mentioned issues with randomization. However, it can be observed that these issues were likely due to random chance, probably because of the block randomization option (Fuhr et al., 2017), since this randomization technique can allocate a group with more secondary illnesses (Kang et al., 2008), exactly as in this case, making data interpretation difficult and introducing biases into the results. Therefore, we disagree with the “low risk of bias” judgment proposed by another recent review (Fuhr et al., 2022).

### 4.1 PRACTICAL IMPLICATIONS

The adoption of hypnosis, or any other intervention, is directly related to the methodological quality of its evidence, as clinical management is based on four pillars: patients, an intervention, a comparison, and the outcomes of interest (Guyatt et al., 2008). This review did not find randomized evidence that currently justifies the clinical use of HT in the treatment of depression.

### 4.2 RESEARCH IMPLICATIONS

These results could justify the proposal and conduct of better clinical trials in HC, as the differences between the biological mechanisms associated with clinical improvement in patients receiving HT and CBT (Haipt et al., 2022), if replicated in confirmatory studies, may be possible evidence of synergy. This could explain the claims that CBT supplemented by hypnosis would be more effective than CBT alone for certain health outcomes (Lynn et al., 2018). Another motivation for conducting better-designed research that could accurately assess the potential enhancement of CBT effectiveness through hypnosis is that patients tend to value emotional and relational aspects more than cognitive ones, which are typically more valued by therapists (Timulak, 2010). Therefore, one possibility to encompass both domains would be the introduction of a clinical tool that could evoke the constructs of feeling and thinking integratively (Alladin & Amundson, 2016).

### 4.3 LIMITATIONS

As with all research, this review also has some important limitations to consider. The literature search revealed the need for a deviation from the protocol (CRD42023409631), which was initially planned to assess the efficacy of HT for MDD. During the literature review phase, it was observed that some of the RCTs from the last 20 years did not focus exclusively on MDD. The heterogeneity of the studies, which included populations with severe depression, depression with anxiety comorbidities, as well as MDD, revealed a significant variation in the characteristics of the participants. This diversity in the study groups compromises the ability to isolate the effects of HT specifically on MDD. In light of this scenario, the scope of the review was expanded to include studies that assessed the efficacy of HT in broader depression conditions, aiming to encompass the variation found and provide a more inclusive and representative analysis of the available research. Another limitation encountered was the impossibility of conducting a meta-analysis as indicated by the main manuals and guidelines in the biomedical literature for interventions due to the absence of RCTs with similar interventions and comparators (Higgins et al., 2022; Kekecs et al., 2022). A considerable limitation to be raised is the sample size of the included studies. It is expected that early studies of an intervention are small and find exaggerated effects, which is why temporal trend analyses in the estimation of a treatment effect are essential (Jiang et al., 2019). However, given that hypnosis is the oldest form of psychotherapy in the West (de Souza et al., 2020), larger RCTs with better methodological quality were to be expected. Another deviation from the protocol was the inclusion of the Cuban RCT (Hernández et al., 2021), as their study accepted participants from the age of 15. However, with the help of another Cuban researcher, we contacted the researchers to ensure that the study population was adult. We decided by consensus to include the study to have a broader scope of the status of HT evidence with larger samples in terms of ethnicities.

This review also has strengths in its favor. To our knowledge, this work features the most comprehensive literature search ever conducted to identify RCTs, not limiting its search by language and also seeking Latin American and Caribbean literature. This review has no authors with potential conflicts of interest regarding the authorship of the included studies, as none of the authors participated in conducting the RCTs analyzed. This work followed the most up-to-date and robust recommendations for conducting its review, using appropriate tools for assessing the risk of bias of individual RCTs (RoB2), evaluating the certainty of evidence (GRADE), and reporting (PRISMA).

## 5. CONCLUSION

There is not enough evidence to indicate that Cognitive Hypnotherapy improves depression severity compared to Cognitive Behavioral Therapy at long-term follow-up (MD -2.10; CI 95% [-6.16, 1.96]; 1 RCT, 84 participants). Based in a single three-arms RCT, there is very-low certainty evidence suggesting that hypnosis plus psychoeducation improves depression severity compared with Psychoeducation alone at medium-term follow-up, with a medium effect size (MD -4.90; CI 95% [-10.05, 0.25]; 27 participants), and very-low certainty evidence suggesting that hypnosis plus psychoeducation it was inferior to Yoga plus psychoeducation improving depression severity with a small effect size at medium-term follow-up (MD 1.00; CI 95% [-3.10, 5.10]; 26 participants). There is not enough evidence to indicate that Hypnotherapy may improve depression severity compared with usual treatment (MD -8.80; CI 95% [-14.96, -2.64]; 1 RCT, 62 participants). Moderate certainty evidence suggests that Hypnotherapy improves depression severity compared with Cognitive Behavioral Therapy at long-term follow up (MD -0.10; CI 95% [-2.99, 2.79]; 1 RCT, 152 participants). There is not enough evidence to indicate that Hypnotherapy improves depression diagnosis rate (remission) compared with Rational emotive behavior therapy (OR 0.20; CI 95% [0.02, 2.02]; 1 RCT; 30 participants). There is not enough evidence to indicate that Mindful Hypnotherapy may improve depression severity compared to wait list (MD -25.56, CI 95% [-30.78, -20.34]; 1 RCT, 31 participants).

### 5.1 AUTHORS CONCLUSION

There is very low-quality evidence suggesting that hypnosis-based interventions may reduce the severity of depression, which prevents the clinical recommendation of this intervention for real-world patients. Better evidence of effectiveness and safety must be produced before such a recommendation can be made, although no significant adverse effects have been found. For hypnotherapy to become a treatment with recognized empirical support, more and better randomized controlled trials need to be conducted to accurately assess the real effect of hypnotherapy in treating depression. Preferably, these trials should include formal diagnoses in the eligibility criteria, pre-publish the analysis plan, and be adequately powered.

## 6. PROTOCOL

This review was pre-registered in PROSPERO and can be accessed with the registration number CRD42023409631.

## 7. FUNDING

This review did not receive any institutional or industrial financial support.

## 8. CONFLICT INTEREST

The lead author of this review has been a hypnotherapist for 7 years and currently works as the clinical director of a psychology clinic in Brazil that also offers hypnotherapy services. The co-supervisor of this work is the editor-in-chief of the International Journal of Clinical and Experimental Hypnosis (IJCEH).

## Data Availability

All data produced in the present work are contained in the manuscript

https://osf.io/639yn/

## 9. ACKNOWLEDGMENTS

F.L.S. - I thank my wife for every day lived by my side. I thank Professor Zoltán Kekecs (ELTE PPK) who warmly welcomed me into his laboratory and provided comments and critiques to improve this work. I also thank my evidence-based practice professors, Igor Eckert, Dr. Leonardo Costa, Dr. Lucíola Costa, and Dr. Eric Pascher.

