## Supplementary material for "Hypnotherapy for major depressive disorder: a systematic review of randomized clinical trials"

Although a formal meta-analysis of the included studies is not feasible due to substantial heterogeneity in the interventions (e.g., HC and MH are different interventions despite being based on hypnosis), populations, and comparators, an exploratory analysis was conducted. This analysis aimed to illustrate the limitations of the current body of evidence and provide a preliminary discussion on potential directions and considerations for future research. It is important to emphasize that the results of this simulated meta-analysis are purely hypothetical and should not be interpreted as direct evidence of the efficacy of hypnosis-based treatments.

**Legend:** 'Simulated' is being used to refer to an analysis with **real data** but purely hypothetical, as a meta-analysis with these studies would be a considerable source of bias.

Instead, they serve as an example to highlight the gaps in existing knowledge and the need for additional well-designed, well-powered, and comparable studies that can be appropriately combined in rigorous meta-analytical analyses capable of demonstrating the true potential effect of HT.

**Supplementary figure 1** - Simulated Meta-Analysis on the Potential Effect of Hypnosis-Based Interventions.

| | Hypnosis | | | Control | | | | Std. Mean Difference |
|---|---|---|---|---|---|---|---|---|
| **Study or Subgroup** | **Mean** | **SD** | **Total** | **Mean** | **SD** | **Total** | **Weight** | **IV, Random, 95% CI** |
| Alladin & Alibhai, 2007 | 13.6 | 10.3 | 42 | 15.7 | 8.6 | 42 | 18.8% | -0.22 [-0.65, 0.21] |
| Butler et al., 2008 | 7.31 | 5.92 | 13 | 6.31 | 5.53 | 13 | 15.7% | 0.17 [-0.60, 0.94] |
| Butler et al., 2008 | 7.31 | 5.92 | 13 | 12.21 | 7.31 | 14 | 15.5% | -0.71 [-1.49, 0.07] |
| Chiu et al., 2018 | 17.07 | 13.14 | 29 | 25.87 | 11.36 | 33 | 18.1% | -0.71 [-1.23, -0.20] |
| Fuhr et al., 2021 | 10.4 | 9.1 | 74 | 10.5 | 9.1 | 78 | 19.7% | -0.01 [-0.33, 0.31] |
| Khazraee et al., 2023 | 8.44 | 7.99 | 16 | 34 | 6.83 | 15 | 12.2% | -3.34 [-4.48, -2.21] |
| **Total (95% CI)** | | | **187** | | | **195** | **100.0%** | **-0.66 [-1.28, -0.05]** |

Heterogeneity: $Tau^2 = 0.48$; $Chi^2 = 35.76$, df = 5 ($P < 0.00001$); $I^2 = 86\%$
Test for overall effect: $Z = 2.11$ ($P = 0.03$)

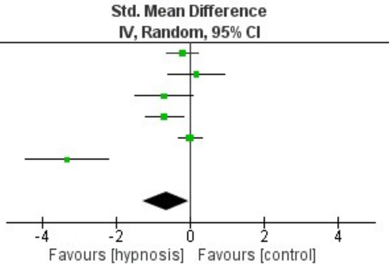


Only with a hypothetical meta-analysis, incorporating data from incomparable RCTs of hypnosis-based interventions from the past 20 years, it was found that HT achieved statistically significant superiority compared to controls (P = 0.03). The simulated meta-analysis indicated a possible moderate effect of hypnosis-based interventions over their controls (SMD -0.66; 95% CI [-1.28, -0.05]). This hypothetical-potential result is only achieved by accounting for the Butler et al. study twice, as the pilot RCT in question had three intervention arms (yoga, hypnosis, and control). When synthesizing the results without double counting (group 1 versus group 2, group 1 versus group 3), the already hypothetical result would also be non-significant compared to controls (SMD -0.66; 95% CI [-1.36, 0.04]) as it touches the null line (Fletcher, 2021). However, we understand that if we had comparable studies, we could perform a multilevel meta-analysis and the Butler study would not necessarily be a source of bias, as would the inclusion of both per-protocol and intention-to-treat analyses (Fuhr et al., 2021), which were not done in this example.

The primary reason this meta-analysis demonstrated a statistically significant difference, suggesting the supposed favorability of hypnosis, is due to the study by Khazraee et al., a study with 31 participants, controlled by a waitlist, which generated an effect size entirely disproportionate to the general medical sciences literature, common in studies with a high risk of bias and small sample sizes (Pereira et al., 2012).

**Supplementary figure 2** - Simulated Meta-Analysis on the Potential Effect of Hypnosis-Based Interventions.

| | Hypnosis | | | Control | | | | Std. Mean Difference |
|---|---|---|---|---|---|---|---|---|
| Study or Subgroup | Mean | SD | Total | Mean | SD | Total | Weight | IV, Random, 95% CI |
| Alladin & Alibhai, 2007 | 13.6 | 10.3 | 42 | 15.7 | 8.6 | 42 | 22.0% | -0.22 [-0.65, 0.21] |
| Butler et al., 2008 | 7.31 | 5.92 | 15 | 6.31 | 5.53 | 15 | 19.2% | 0.17 [-0.55, 0.89] |
| Chiu et al., 2018 | 17.07 | 13.14 | 29 | 25.87 | 11.36 | 33 | 21.2% | -0.71 [-1.23, -0.20] |
| Fuhr et al., 2021 | 10.4 | 9.1 | 74 | 10.5 | 9.1 | 78 | 22.9% | -0.01 [-0.33, 0.31] |
| Khazraee et al., 2023 | 8.44 | 7.99 | 16 | 34 | 6.83 | 15 | 14.7% | -3.34 [-4.48, -2.21] |
| **Total (95% CI)** | | | **176** | | | **183** | **100.0%** | **-0.66 [-1.36, 0.04]** |

Heterogeneity: Tau² = 0.53; Chi² = 34.95, df = 4 (P < 0.00001); I² = 89%
Test for overall effect: Z = 1.85 (P = 0.06)

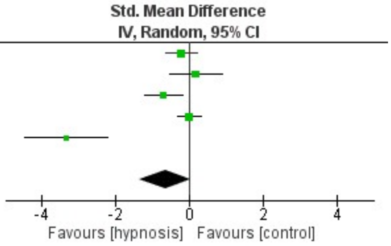


As discussed earlier in this document, if we remove the artificial addition of 13 patients from the double-counting of the hypnosis group in Butler et al. (figure 2), the difference becomes non-significant, though by a small margin, due to the impact of the Khazraee et al. study.

Overall, this document aims to clarify potential doubts of readers regarding the effects of hypnosis-based interventions for depression. Our systematic review did not find randomized evidence to justify its clinical use until more and better research is conducted, and **even a simulated meta-analysis, which would be considered biased, also did not find evidence that would justify its recommendation**.
